# Post-birth exposure contrasts for children during the Household Air Pollution Intervention Network randomized controlled trial

**DOI:** 10.1101/2023.07.04.23292226

**Authors:** Ajay Pillarisetti, Wenlu Ye, Kalpana Balakrishnan, Ghislaine Rosa, Anaité Díaz-Artiga, Lindsay J. Underhill, Kyle Steenland, Jennifer L. Peel, Miles A. Kirby, John McCracken, Lance Waller, Howard Chang, Jiantong Wang, Ephrem Dusabimana, Florien Ndagijimana, Sankar Sambandam, Krishnendu Mukhopadhyay, Katherine A. Kearns, Devan Campbell, Jacob Kremer, Joshua Rosenthal, Ahana Ghosh, Maggie Clark, William Checkley, Thomas Clasen, Luke Naeher, Ricardo Piedrahita, Michael Johnson, Household Air Pollution Intervention Network (HAPIN) trial Investigators

## Abstract

Exposure to household air pollution is a leading cause of ill-health globally. The Household Air Pollution Intervention Network (HAPIN) randomized controlled trial evaluated the impact of a free liquefied petroleum gas stove and fuel intervention on birth outcomes and maternal and child health. As part of HAPIN, an extensive exposure assessment was conducted. Here, we report on PM_2.5_ and CO exposures of young children (≤ 15 months old) reconstructed using a Bluetooth-beacon based time-activity monitoring system coupled with microenvironmental pollutant monitors. Median (IQR) exposures to PM_2.5_ were 65.1 (33 – 128.2) µg/m^3^ in the control group and 22.9 (17.2 – 35.3) µg/m3 in the intervention group; for CO, median (IQR) exposures were 1.1 (0.3 – 2.9) ppm and 0.2 (0 – 0.7) ppm for control and intervention group, respectively. Exposure reductions were stable over time and consistent with previous findings for the children’s mothers. In the intervention group, 75% of children’s reconstructed exposures were below the WHO interim target guideline value of 35 µg/m^3^, while 26% were below the standard in the control group. Our findings suggest that an LPG fuel and stove intervention can substantially reduce children’s exposure to household air pollution.

## Introduction

Between 3 and 4 billion people cook with solid fuels (coal, wood, dung, or crop residues)^1, 2^, resulting in high exposures to household air pollution (HAP), including particulate matter with an aerodynamic diameter of 2.5 µm or less (PM_2.5_), carbon monoxide (CO), and black carbon (BC), among others. In 2019, 2.3 million deaths were attributed to HAP exposure, approximately 460,000 of which occurred in children under the age of 5.^3^

HAP-reducing interventions – including improved or advanced biomass stoves and clean fuel stoves – have had varying exposure reduction effectiveness to date.^4–6^ Trials evaluating the impact of interventions on health have often insufficiently measured changes in exposures^7^ or failed to reduce exposure to levels expected to result in improved health.^8, 9^ Among those that do measure exposure, many have reported little or no change in exposure, arguably due to mixed use of intervention and traditional stoves or poor-performing interventions that do not reduce exposure in real-world settings.^10, 11^

The multi-country Household Air Pollution Intervention Network (HAPIN) clean stove and fuel randomized controlled trial (RCT) sought to address these shortcomings. First, it provided a liquefied petroleum gas LPG stove and continuous clean fuel supply to pregnant women, with behavioral reinforcement, leading to high fidelity and adherence to the intervention during pregnancy^12^ and after childbirth.^13^ Second, we performed extensive personal exposure assessments among participants throughout the trial. Among expectant mothers, we found significantly lower exposures to HAP among the intervention group than in the control group: PM_2.5_ exposures were 66% lower (23.9 vs. 70.7 µg/m^3^), BC exposures were 71% lower (2.8 vs. 9.6 µg/m^3^), and CO exposures were 83% lower (0.2 vs. 1.1 ppm).^14^

Exposures are often measured for primary cooks, typically women; less is known about exposures to infants and young children,^15^ despite evidence from the ambient air pollution literature that early-in-life exposure can have respiratory^16^ and cardiovascular^17, 18^ impacts that may set health trajectories into adulthood. The scarcity of exposure data on children is due partly to challenges of wearability, compliance, and induced behavior change; alternatively, exposures can be reconstructed using microenvironmental measurements coupled with time-activity data either from sensors^19, 20^ or self-reported questionnaires.^21^ Here, we report the impact of HAPIN’s LPG intervention on estimated PM_2.5_ and CO exposures for HAPIN children using indirect exposure reconstruction methods validated during HAPIN’s formative phase.^20^

## Materials and Methods

### Study population

The HAPIN RCT of an LPG fuel and stove intervention occurred in Guatemala, India, Peru, and Rwanda. The approach and sites have been described in detail.^22–24^ Briefly, we selected rural sites in each country with low background air pollution concentrations, few additional air pollution sources, and where most households used biomass stoves for cooking.^14, 25, 26^ The trial assessed intervention health effects among pregnant women (n ∼ 3200), their resulting newborn children (n ∼ 3200), and non-pregnant adult women living in the same household (n ∼ 418), split evenly between intervention and control arms. Primary health outcomes are birth weight,^27, 28^ incidence of severe pediatric pneumonia,^29^ stunted growth in the children,^22^ and blood pressure in the non-pregnant adult women.^22^

The study protocol has been reviewed and approved by institutional review boards (IRBs) and Ethics Committees at Emory University (00089799), Johns Hopkins University (00007403), Sri Ramachandra Institute of Higher Education and Research (IEC-N1/16/JUL/54/49), the Indian Council of Medical Research – Health Ministry Screening Committee (5/8/4-30/(Env)/Indo-US/2016-NCD-I), Universidad del Valle de Guatemala (146-08-2016), Guatemalan Ministry of Health National Ethics Committee (11-2016), Asociación Benefica PRISMA (CE2981.17), the London School of Hygiene and Tropical Medicine (11664-5), the Rwandan National Ethics Committee (No.357/RNEC/2018), and Washington University in St. Louis (201611159). The study has been registered with ClinicalTrials.gov (Identifier NCT02944682).

### Recruitment

Pregnant women were identified and enrolled through partnerships with local clinics and community health workers. Eligible women were aged 18-35 years old at 9 to <20 weeks’ gestation, with a viable singleton pregnancy (confirmed by ultrasound); used primarily biomass fuel for cooking; and agreed to participate via informed consent. Exclusion criteria included tobacco use, plans to move outside the study area, and plans to switch to primary use of clean fuels.

### Intervention

Participants were randomly assigned (one-to-one) to receive an LPG stove, continuous fuel delivery, and regular behavioral messaging versus continued use of a biomass-burning stove. In India and Peru, stratified randomization was used to ensure balance between distinct geographical regions within each country. No additional stratification was used in Rwanda and Guatemala, where the study areas were deemed homogenous.

The HAPIN intervention was refined during formative research and described in detail previously.^22^ Stoves and fuel were procured at each IRC based on local cooking practices and equipment and fuel availability. The exact stove deployed varied by country; all stoves had at least two burners and included additional components for preparation of traditional foods (e.g., a flat griddle for cooking tortillas in Guatemala; a roasting device in Rwanda). Intervention households received the stove and a continuous supply of LPG at no cost throughout follow-up. Behavioral support included (i) a pledge by intervention homes to use the LPG stove for all cooking throughout the trial, (ii) safety training, (iii) tailored messaging to encourage exclusive use of LPG and discourage use of traditional stoves, and (iv) behavioral reinforcements upon detection of traditional stove use.

### Monitoring plan and microenvironmental PM_2.5_ and CO measurements

We attempted collection of three 24-hour PM_2.5_ and CO measurements in children in the first year of life (<3 months, ∼6 months, and ∼12 months). We used a validated indirect approach to assess the exposure of children; using standard personal exposure monitors is impractical due to their size and participant preferences.^22^ This approach pairs microenvironmental pollutant sampling and participant proximity sensing.^23^ Details of the indirect exposure assessment process and approach are described below. Microenvironmental PM_2.5_ concentrations were measured using the Enhanced Children’s MicroPEM (ECM, RTI International, Durham, NC USA), the same device used for personal PM_2.5_ exposure monitoring in the HAPIN trial. The ECM measures PM_2.5_ concentrations every 10 seconds using a nephelometer and simultaneously collects integrated gravimetric samples on 15 mm polytetrafluoroethylene filters (Measurement Technology Laboratories).^23^ Gravimetrically corrected nephelometric PM_2.5_ concentrations are used in infants’ indirect PM_2.5_ exposure estimates. Continuous microenvironmental CO concentrations at 1-minute intervals were logged using the Lascar EL-USB-300 (Lascar Electronics), a large pen-sized device with a sensing range between 0 and 300 ppm. Details on instrumentation, sampling strategy, sample analysis, data process, and quality have been described previously.^14, 23^

### Indirect exposure measurements

Infants’ exposure is estimated using an indirect microenvironmental approach linking objectively determined locations (microenvironments) and assessed PM_2.5_/CO concentrations in these same areas at the times when the infant was deemed present in the microenvironment with the monitor. We used a Bluetooth^®^-based beacon system for time-location monitoring among infants. The beacon system consists of 1) emitters (Model O, Roximity Inc. Denver, CO, USA or EM Microelectronic, La Tène, Neuchâtel, Switzerland) and 2) loggers (Berkeley Air Monitoring Group, Berkeley, CA, USA) to monitor the location of infants within a home. Monitored microenvironments include the most commonly occupied rooms (i.e., the main living/sleeping areas, the kitchen), and mothers who consented to be a mobile microenvironment and wore an exposure monitor and beacon logger.^23^

The emitter (hereafter referred to as a beacon) is a coin-size device that constantly emits Bluetooth signals.^20^ Infants wore two Beacons each on their clothing during the sampling period. The Beacon logger is a smartphone-sized device that receives and logs Bluetooth signals emitted from the Beacon.^20^ The beacon logger records the beacon’s unique media access control (MAC) address and the received signal strength indicator (RSSI) every 20 seconds. The RSSI value is proportional to the distance between the Beacon and the Beacon logger and is thus used to determine the infant’s microenvironmental location.^20^

In each microenvironment, we co-located a fixed-position logger with a PM_2.5_ and a CO monitor. The time-microenvironment and corresponding area concentrations are assigned at 5-minute intervals as the Beacon logger recorded the strongest average RSSI from the two Beacons worn by infants.^20^ Reconstructed personal PM_2.5_/CO exposures for infants were estimated by integrating corresponding microenvironmental concentrations with time spent in the respective locations within the 24-hour monitoring period.

### Statistical analysis

All analyses were performed in R (version 4). We calculated descriptive statistics for valid infant indirect PM_2.5_ and CO exposures by study group (control vs. intervention), study visit (<3 months, ∼6 months, and ∼12 months), and IRC. We evaluated the overall and stove/fuel stratified Spearman’s correlations 1) of measurements of the same pollutants collected at each visit and 2) between PM_2.5_ and CO exposure at each measurement point. We assessed the differences in PM_2.5_ and CO exposures between the controls and interventions by study visit (at <3 months, ∼6 months, and ∼12 months) using non-parametric tests (Wilcoxon Rank Sum, Kruskal-Wallis, and Dunn’s tests). Moreover, we evaluated the proportion of children’s exposures that were less than or equal to the WHO Annual Interim Target 1 (WHO IT-1) level of 35 μg/m^3^ for PM_2.5_ and the WHO 24-hour guideline value of 4 mg/m^3^ (∼3.5 ppm) for CO. Additionally, we visually examined the trends in PM_2.5_ and CO exposures by plotting exposures against the age of infants.

Since each child had up to three exposure measurements, we used mixed-effects models to assess the impact of the intervention on personal exposures. We included a random intercept to account for correlations among repeated measurements on the same child. Exposures are natural log-transformed in the models given their right-skewed distribution. As secondary analyses, we also modeled non-transformed exposures to assess the absolute changes in exposures. The model (**Eq. 1**) assesses the difference in mean PM2.5/CO exposures between the control and intervention groups during the first year of life.

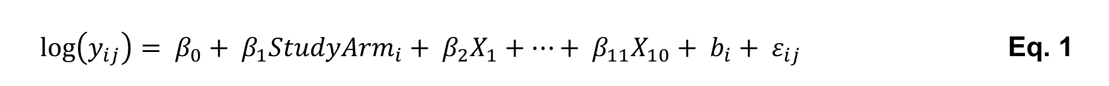

where *y*_*ij*_ is the PM_2.5_ or CO exposure for child *i* at visit *j* (<3 months, ∼6 months, or ∼12 months visit), *Study Arm_j_* is the child’s assigned study group (reference: control), *X*_$_ through *X*_$#_ are indicator variables for the study sites within which the randomization took place (1 in Guatemala, 2 in India, 6 in Peru, and 1 in Rwanda), *b*_*i*_ is the random intercept for child *i*, *e*_*ij*_ represents the model residual, assumed to be normally distributed. The parameter of interest (Study Arm) was exponentiated, subtracted from one, and multiplied by 100 to obtain the percent reduction in personal exposure due to intervention. In addition, we calculated the intraclass correlation coefficient (ICC) using mixed-effects models with a random intercept for each child to evaluate the within and between individual variability.

## Results and Discussion

*Household characteristics.* 3195 pregnant women randomized to the control (N = 1605) or intervention (N = 1590) groups yielded 3061 live births (control: 1525; intervention: 1536). Among those, 75 exited the study post-birth, and 2986 children remained in the trial (control: 1489; intervention: 1497). **Table 1** summarizes selected baseline household and child characteristics by study group and IRC. The household characteristics of children who remained in the trial were similar between the control and intervention groups at baseline trial-wide and within each IRC. About 80% of households had access to electricity at home, which was also reported as the primary light source across the four IRCs. The primary fuel type varied in each IRC, with wood being predominant in Guatemala and India. Cow dung was reported as the primary fuel in Peru. In Rwanda, most households used wood, but a quarter reported charcoal as the primary household energy source. Overall, 47% of the households owned only one stove (mainly in India and Rwanda), and 48% used two stoves (mainly in Guatemala and Peru). Trial-wide only 10% of the households reported having one or more active smokers at home, mainly from India (34%). The mean (SD) gestational age at delivery was 39.3 (1.5) weeks for the control group and 39.4 (1.5) weeks for the intervention group. 4.8% (N = 71) and 5.1% (N = 77) of the births were classified as preterm (defined as gestational age at birth < 37 weeks) in the control and intervention groups, respectively.

**Table 1.** Household and other adult women participant characteristics at baseline, by IRC and study arm.

| Variable | Guatemala |  | India |  | Peru |  | Rwanda |  | Trial-wide |  |
| --- | --- | --- | --- | --- | --- | --- | --- | --- | --- | --- |
|  | Control<br>(n = 378) | Intervention<br>(n = 376) | Control<br>(n = 380) | Intervention<br>(n = 381) | Control<br>(n = 347) | Intervention<br>(n = 375) | Control<br>(n = 384) | Intervention<br>(n = 365) | Control<br>(n = 1489) | Intervention<br>(n = 1497) |
| *Household and kitchen characteristics* |  |  |  |  |  |  |  |  |  |  |
| **Household Size** |  |  |  |  |  |  |  |  |  |  |
| Mean (SD) | 5.1 (2.6) | 5.2 (2.6) | 3.8 (1.5) | 3.8 (1.6) | 4.6 (1.7) | 4.5 (1.8) | 3.5 (1.5) | 3.5 (1.5) | 4.3 (2) | 4.3 (2) |
| Range | 2-18 | 2-13 | 2-9 | 1-10 | 2-10 | 2-11 | 1-10 | 1-10 | 1-18 | 1-13 |
| **Access to electricity, n (%)** |  |  |  |  |  |  |  |  |  |  |
| No | 35 (9.3) | 42 (11.2) | 15 (3.9) | 12 (3.1) | 20 (5.8) | 20 (5.3) | 259 (67.4) | 202 (55.3) | 329 (22.1) | 276 (18.4) |
| Yes | 342 (90.7) | 334 (88.8) | 365 (96.1) | 369 (96.9) | 327 (94.2) | 355 (94.7) | 99 (25.8) | 142 (38.9) | 1134 (76.2) | 1200 (80.2) |
| Missing | 0 | 0 | 0 | 0 | 0 | 0 | 26 (6.8) | 21 (5.8) | 26 (1.7) | 21 (1.4) |
| **Kitchen volume (m3)** |  |  |  |  |  |  |  |  |  |  |
| Mean (SD) | 33.3 (16.2) | 34.1 (17.4) | 20.5 (13.8) | 22 (16.1) | 19.9 (12.7) | 18.9 (11.7) | 11.7 (6.3) | 13.2 (7.1) | 22.2 (15.3) | 23.0 (16.1) |
| Range | 7.1-99.9 | 6.9-125 | 0.4-86.2 | 0.6-135 | 1.8-117 | 0.1-83 | 0.2-41.5 | 0.2-52.5 | 0.2-117 | 0.1-135 |
| Missing, n (%) | 21 (5.6) | 21 (5.6) | 12 (3.2) | 8 (2.1) | 94 (27.1) | 95 (25.3) | 125 (32.6) | 114 (31.2) | 252 (51.4) | 238 (48.6) |
| **Roof in the kitchen, n (%)** |  |  |  |  |  |  |  |  |  |  |
| No | 1 (0.3) | 1 (0.3) | 1 (0.3) | 3 (0.8) | 76 (21.9) | 82 (21.9) | 117 (30.5) | 110 (30.1) | 195 (13.1) | 196 (13.1) |
| Yes | 373 (98.7) | 374 (99.5) | 379 (99.7) | 378 (99.2) | 269 (77.5) | 293 (78.1) | 266 (69.3) | 254 (69.6) | 1287 (86.4) | 1302 (86.8) |
| Missing, n (%) | 4 (1.1) | 1 (0.3) | 0 | 0 | 2 (0.6) | 0 | 1 (0.3) | 1 (0.3) | 7 (0.5) | 2 (0.1) |
| **Number of stoves, n (%)** |  |  |  |  |  |  |  |  |  |  |
| One | 132 (34.9) | 123 (32.7) | 259 (68.2) | 268 (70.3) | 92 (26.5) | 88 (23.5) | 235 (61.4) | 213 (58.4) | 718 (48.3) | 692 (46.3) |
| Two | 216 (57.1) | 219 (58.2) | 118 (31.1) | 107 (28.1) | 245 (70.6) | 276 (73.8) | 129 (33.7) | 136 (37.3) | 708 (47.6) | 738 (49.3) |
| Three or more | 30 (7.9) | 34 (9.0) | 3 (0.8) | 6 (1.6) | 9 (2.6) | 9 (2.4) | 18 (4.7) | 15 (4.1) | 60 (4.0) | 64 (4.3) |
| Missing | 0 | 0 | 0 | 0 | 1 (0.3) | 1 (0.3) | 1 (0.3) | 1 (0.3) | 2 (0.1) | 2 (0.1) |
| **Primary stove has a chimney, n (%)** |  |  |  |  |  |  |  |  |  |  |
| No | 294 (77.8) | 296 (78.7) | 378 (99.5) | 379 (99.5) | 216 (62.2) | 237 (63.2) | 363 (94.5) | 353 (96.7) | 1251 (84.0) | 1265 (84.5) |
| Yes | 84 (22.2) | 80 (21.3) | 2 (0.5) | 2 (0.5) | 130 (37.5) | 138 (36.8) | 20 (5.2) | 11 (3.0) | 236 (15.8) | 231 (15.4) |
| Missing | 0 | 0 | 0 | 0 | 1 (0.3) | 0 | 1 (0.3) | 1 (0.3) | 2 (0.1) | 1 (0.1) |
| **Primary fuel type, n (%)** |  |  |  |  |  |  |  |  |  |  |
| Cow dung | 0 | 0 | 0 | 0 | 303 (87.3) | 328 (87.5) | 0 | 0 | 303 (20.3) | 328 (21.9) |
| Wood | 3712(98.4) | 375 (99.7) | 380 (100%) | 381 (100%) | 41 (11.8) | 39 (10.4) | 305 (79.4) | 238 (65.2) | 1098 (73.7) | 1033 (69.0) |
| Charcoal | 0 | 0 | 0 | 0 | 0 | 0 | 70 (18.2) | 116 (31.8) | 70 (4.7) | 116 (7.7) |
| Other | 2 (0.5) | 1 (0.3) | 0 | 0 | 2 (0.6) | 8 (2.1) | 8 (2.1) | 10 (2.7) | 12 (0.8) | 19 (1.3) |
| Missing | 4 (1.1) | 0 | 0 | 0 | 1 (0.3) | 0 | 1 (0.3) | 1 (0.3) | 6 (0.4) | 1 (0.1) |
| **Primary stove location, n (%)** |  |  |  |  |  |  |  |  |  |  |
| In participant's bedroom | 17 (4.5) | 17 (4.5) | 70 (18.4) | 93 (24.4) | 0 | 4 (1.1) | 2 (0.5) | 4 (1.1) | 89 (6.0) | 118 (7.9) |
| Room immediately adjacent to the participant's bedroom | 189 (50.0) | 173 (46.0) | 140 (36.8) | 115 (30.2) | 27 (7.8) | 22 (5.9) | 9 (2.3) | 14 (3.8) | 365 (24.5) | 324 (21.6) |
| Separated from the participant's bedroom but inside the house | 97 (25.7) | 115 (30.6) | 67 (17.6) | 67 (17.6) | 56 (16.1) | 69 (18.4) | 18 (4.7) | 19 (5.2) | 238 (16.0) | 270 (18.0) |
| Outside the house (outdoors) | 5 (1.3) | 7 (1.9) | 1 (0.3) | 2 (0.5) | 85 (24.5) | 87 (23.2) | 117 (30.5) | 115 (31.5) | 208 (14.0) | 211 (14.1) |
| In a separate building detached from the bedroom-main home | 69 (18.3) | 64 (17.0) | 102 (26.8) | 104 (27.3) | 177 (51.0) | 192 (51.2) | 237 (61.7) | 212 (58.1) | 585 (39.3) | 572 (38.2) |
| Other/Missing | 1 (0.3) | 0 | 0 | 0 | 2 (0.6) | 1 (0.3) | 1 (0.3) | 1 (0.3) | 4 (0.3) | 2 (0.1) |
| **Primary light source, n (%)** |  |  |  |  |  |  |  |  |  |  |
| Torch (battery) | 5 (1.3) | 5 (1.3) | 4 (1.1) | 7 (1.8) | 4 (1.2) | 7 (1.9) | 84 (21.9) | 79 (21.6) | 97 (6.5) | 98 (6.5) |
| Kerosene lamp | 1 (0.3) | 5 (1.3) | 8 (2.1) | 7 (1.8) | 1 (0.3) | 0 | 36 (9.4) | 20 (5.5) | 46 (3.1) | 32 (2.1) |
| Solar light | 0 | 2 (0.5) | 2 (0.5) | 1 (0.3) | 10 (2.9) | 11 (2.9) | 132 (34.4) | 108 (29.6) | 144 (9.7) | 122 (8.1) |
| Electricity | 343 (90.7) | 328 (87.2) | 366 (96.3) | 366 (96.1) | 319 (91.9) | 342 (91.2) | 83 (21.6) | 127 (34.8) | 1111 (74.6) | 1163 (77.7) |
| Other | 29 (7.7) | 36 (9.6) | 0 | 0 | 12 (3.5) | 15 (4.0) | 48 (12.5) | 30 (8.2) | 89 (6.0) | 81 (5.4) |
| Missing | 0 | 0 | 0 | 0 | 1 (0.3) | 0 | 1 (0.3) | 1 (0.3) | 2 (0.1) | 1 (0.1) |
| **Presence of a smoker in home, n (%)** |  |  |  |  |  |  |  |  |  |  |
| No | 358 (94.7) | 357 (94.9) | 252 (66.3) | 268 (70.3) | 343 (98.8) | 372 (99.2) | 365 (95.1) | 357 (97.8) | 1318 (88.5) | 1354 (90.4) |
| Yes | 20 (5.3) | 19 (5.1) | 128 (33.7) | 113 (29.7) | 3 (0.9) | 3 (0.8) | 18 (4.7) | 7 (1.9) | 169 (11.3) | 142 (9.5) |
| Missing | 0 | 0 | 0 | 0 | 1 (0.3) | 0 | 1 (0.3) | 1 (0.3) | 2 (0.1) | 1 (0.1) |
| *Child characteristics* |  |  |  |  |  |  |  |  |  |  |
| **Child sex, n (%)** |  |  |  |  |  |  |  |  |  |  |
| Female | 186 (49.2) | 178 (47.3) | 174 (45.8) | 177 (46.5) | 176 (50.7) | 185 (49.3) | 183 (47.7) | 181 (49.6) | 719 (48.3) | 721 (48.2) |
| Male | 192 (50.8) | 198 (52.7) | 206 (54.2) | 204 (53.5) | 171 (49.3) | 190 (50.7) | 201 (52.3) | 184 (50.4) | 770 (51.7) | 776 (51.8) |
| **Gestational age at delivery, n (%)** |  |  |  |  |  |  |  |  |  |  |
| Mean (SD) | 39.2 (1.3) | 39.2 (1.5) | 38.9 (1.4) | 38.9 (1.5) | 39.3 (1.4) | 39.4 (1.3) | 40 (1.6) | 39.9 (1.6) | 39.3 (1.5) | 39.4 (1.5) |
| Range | 31.7-42.6 | 32.6-43.4 | 32.3-41.7 | 29.6-42.3 | 29.4-43.3 | 33.6-42.6 | 30.6-44.4 | 29.0-44.0 | 29.4-44.4 | 29.0-44.0 |
| **Preterm birth, n (%)** |  |  |  |  |  |  |  |  |  |  |
| No | 356 (94.2) | 355 (92.4) | 358 (94.2) | 355 (93.2) | 336 (96.8) | 357 (95.2) | 368 (95.8) | 353 (96.7) | 1419 (95.2) | 1420 (94.9) |
| Yes | 22 (5.8) | 21 (5.6) | 22 (5.8) | 26 (6.8) | 11 (3.2) | 18 (4.8) | 16 (4.2) | 12 (3.3) | 71 (4.8) | 77 (5.1) |

### Indirect exposure measurements, data completeness, compliance, and quality assurance and control

HAPIN field staff made 7227 exposure monitoring visits for 2986 children during their first year of life. 61% and 63% of the children had at least one valid indirect PM_2.5_ and CO exposure sample, respectively. The proportion of (a) enrolled children with valid indirect measurements and (b) exposure visits with valid indirect measurements vary by study visit and IRC (**Table S1-S4**). Generally, the percentage of valid samples went down over time; the <3 month visit had the most valid samples, and the ∼12 month visit had the least. The moderate to low percentage of valid samples is partially due to increased equipment failure as time passed, field measurements interrupted by the COVID-19 pandemic lockdown, and the challenge and complexity of successfully merging multiple real-time data sources in the indirect microenvironmental approach.

### Child exposure summary

24-hour child indirect exposures to PM_2.5_ and CO are summarized by study arm and visit in **Table 2** and in **Figure 1**. Exposure visits occurred as planned, with the first visit occurring on average 3.2 (SD 0.43) months after birth, the second visit occurring on average 6.3 (SD 0.48) months after birth, and the final visit occurring 12.1 (0.49) months after birth (**SI Figure S1**).

**Table 2.** Summary of 24-hour child indirect exposure to PM_2.5_ and CO by study arm and visit.

| Visit |  | PM<sub>2.5</sub> Exposure |  | CO Exposure |  |
| --- | --- | --- | --- | --- | --- |
|  |  | Control | Intervention | Control | Intervention |
| B1 | N | 629 | 554 | 604 | 620 |
|  | Mean (SD) | 108.8 (125) | 39.3 (119.5) | 2.4 (4) | 0.8 (2.3) |
|  | Median (IQR) | 66.5 (33.7 — 123.7) | 23.1 (17.4 — 36.9) | 1.1 (0.3 — 2.8) | 0.2 (0 — 0.6) |
|  | Range | 9.6-1037.7 | 0.2-2294 | 0-43.1 | 0-40.9 |
| B2 | N | 465 | 452 | 459 | 516 |
|  | Mean (SD) | 111.9 (146.9) | 34.6 (69.4) | 2.6 (4.6) | 0.7 (1.5) |
|  | Median (IQR) | 59.9 (30.8 — 122.7) | 23 (17.1 — 34.7) | 1.1 (0.3 — 3.1) | 0.2 (0 — 0.7) |
|  | Range | 8.6-1133.2 | 1.1-1068.8 | 0-51.8 | 0-16.5 |
| B4 | N | 358 | 318 | 341 | 366 |
|  | Mean (SD) | 123.6 (161.6) | 38.6 (87.5) | 2.7 (5.1) | 0.9 (2.6) |
|  | Median (IQR) | 71.8 (35.8 — 139) | 22 (17 — 33.6) | 1 (0.3 — 2.7) | 0.1 (0 — 0.7) |
|  | Range | 9.4-1393.9 | 6.9-1121 | 0-58.2 | 0-31.6 |
| Post-birth<br>Overall | N | 1452 | 1324 | 1404 | 1502 |
|  | Mean (SD) | 113.4 (141.9) | 37.5 (97.2) | 2.5 (4.5) | 0.8 (2.1) |
|  | Median (IQR) | 65.1 (33 — 128.2) | 22.9 (17.2 — 35.3) | 1.1 (0.3 — 2.9) | 0.2 (0 — 0.7) |
|  | Range | 8.6-1393.9 | 0.2-2294 | 0-58.2 | 0-40.9 |
| Post-birth<br>Overall<br>(household-<br>average<br>measures) | N | 933 | 848 | 916 | 965 |
|  | Mean (SD) | 109.3 (122.7) | 37.8 (97.4) | 2.6 (4.1) | 0.9 (2) |
|  | Median (IQR) | 66 (35.2 — 132.2) | 24.2 (17.8 — 36.4) | 1.3 (0.4 — 3) | 0.3 (0 — 0.8) |
|  | Range | 8.6-832.5 | 0.2-2294 | 0-51.8 | 0-31.6 |

**Figure 1.**
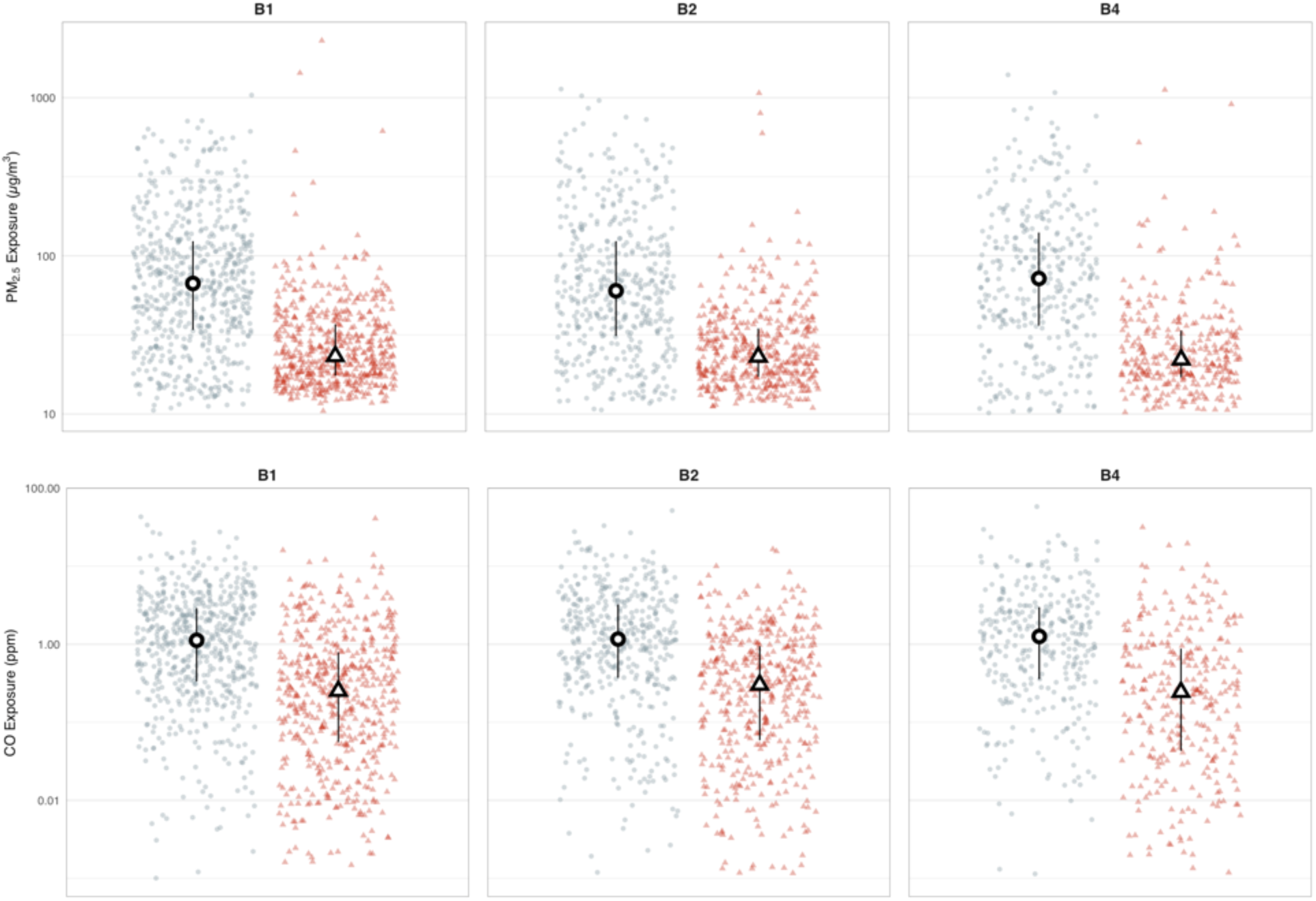
24-hour child indirect PM_2.5_ (upper) and CO (lower) exposures. Red (right) and blue (left) dots are study round samples in intervention and control households, respectively. Circles and triangles outlined in black are median values in control and intervention households, respectively. Lines are interquartile ranges. B1 = post-birth visit 1 (< 3 months, N_control_ = 584, N_intervention_ = 531), B2 = post-birth visit 2 (∼ 6 months, N_control_ = 441, N_intervention_ = 444), B3 = post-birth visit 3 (∼ 12 months, N_control_ = 325, N_intervention_ = 300)

Trial-wide and within IRCs, we observed stable reductions in PM_2.5_ between post-birth measurement rounds. The overall median (IQR) child PM_2.5_ and CO exposures are 65.1 (33.0 — 128.2) µg/m^3^ and 1.1 (0.3 — 2.9) ppm for the control group, and 22.9 (17.2 — 35.3) µg/m^3^ and 0.2 (0.0 — 0.7) ppm for the intervention group. Median child exposures to PM_2.5_ in the intervention arm were 65% lower than in the control arm during the <3 months visit (67 vs. 23 µg/m^3^), 62% lower at the ∼6 months visit (60 vs. 23 µg/m^3^), and 69% lower at the ∼12 months visit (72 vs. 22 µg/m^3^). CO exposures were similarly reduced: 85% at <3 months (1.1 vs. 0.2 ppm), 82% at ∼6 months (1.1 vs. 0.2 ppm), and 88% at ∼12 months (1.0 vs 0.1 ppm). Across all rounds, 26% of control samples and 75% of intervention samples were below the WHO-IT1 target guideline value of 35 µg/m^3^. Similarly, 95% of intervention samples and 79% of control samples were below the WHO annual guideline value for CO (3.5 ppm). Both descriptive statistics of PM_2.5_ and CO exposures and the proportion of measurements below the WHO guidelines were similar across the three post-birth visits.

### Exposures over time

We plotted children’s exposures to PM_2.5_ by age (the number of days between the exposure measurement date and the child’s birthday, **Figure 2**). We observe consistent and distinct separation of exposures between the control and intervention children in the first year of life. Control and intervention samples were significantly different at each round for each pollutant (p < 0.001). For control households, there was no significant difference between PM_2.5_ or CO exposures by study visits. Exposures were 109 (SD 125), 112 (SD 147), and 124 (SD 162) for <3 months, ∼6 months, and ∼12 months visits. Similar trends were observed for CO: 2.4 (SD 4.0), 2.6 (SD 4.6), and 2.7 (SD 5.1) ppm at < 3 months, ∼6 months, and ∼ 2 months. There were no significant differences between PM_2.5_ or CO exposures by study round for intervention households. Exposures were 39 (SD 120), 35 (SD 69), and 39 (SD 88) for visits at <3 months, ∼6 months, and ∼12 months. Similar trends were observed for CO: 0.8 (SD 2.3), 0.7 (SD 1.5), and 0.9 (SD 2.6) ppm at <3 months, ∼6 months, and ∼12 months.

**Figure 2.**
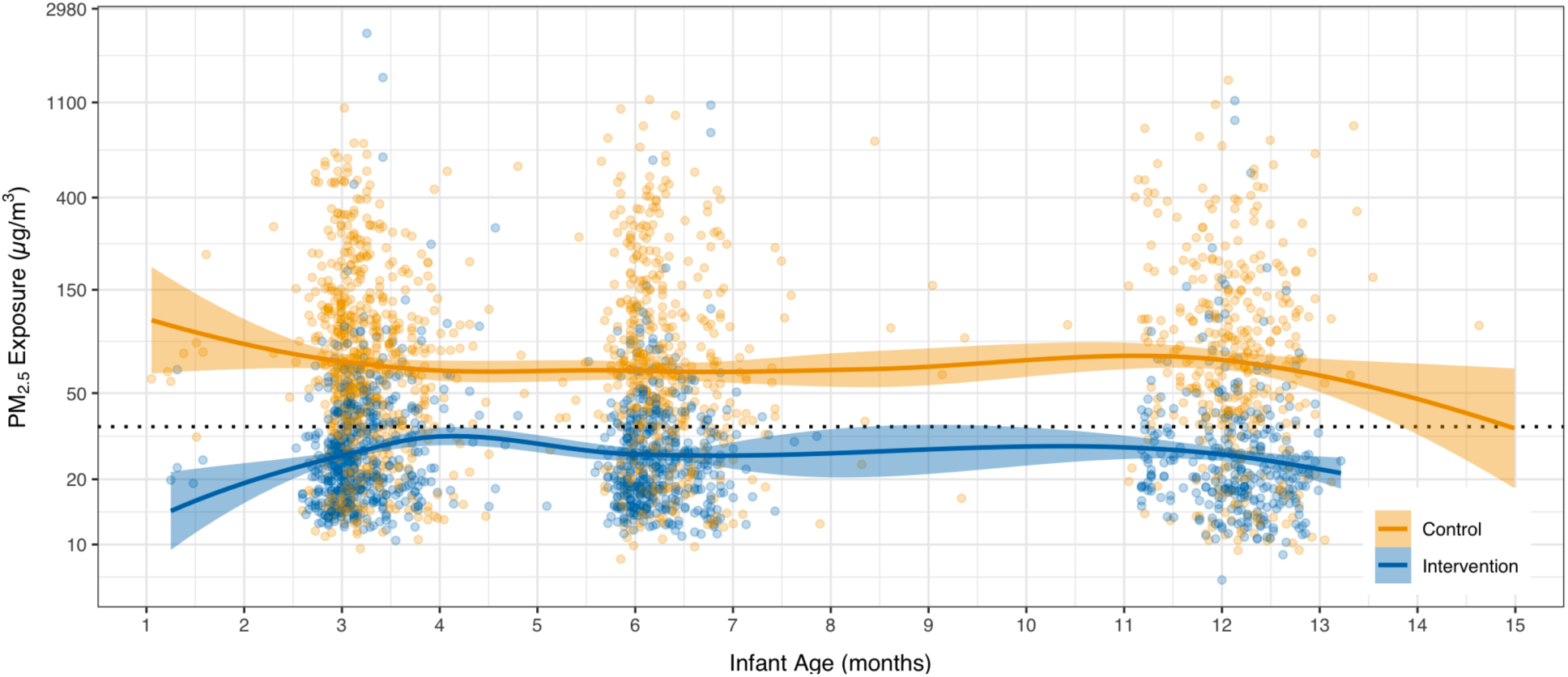
Trends in PM_2.5_ exposure. The x-axis is the infant age in days, calculated as the number of days between the exposure measurement date and a child’s birthday. Solid lines are a locally weighted smoothing (LOESS) function. Shaded areas are standard errors. Yellow (lighter) points are individual data points from control households; blue (darker) points are from intervention households.

### Correlations between pollutants and between study rounds

Correlations between pollutants were moderate for the control arm (Spearman’s ρ 0.45) and poor for the intervention arm (Spearman’s ρ 0.08). Overall correlations between study rounds (depicted in **SI Figure S2**) were weak for CO (Spearman’s ρ 0.37) and were moderate for PM (Spearman’s ρ 0.58); for both pollutants, they were stronger in the control arm than in the intervention arm.

### Modeling the effect of the LPG stove and fuel intervention on personal exposures

Modeled impacts of the HAPIN LPG fuel and stove intervention on children’s exposure indicated significant reductions for PM_2.5_ and CO (**Figure 3**) of 62% (95% CI 59, 64) and 80 (95% CI 77, 83), respectively. In models with untransformed outcomes, these percent reductions translate to predicted absolute reductions of 79.9 µg/m^3^ (95% CI 69.7, 90.1) and 1.8 ppm (95% CI 1.5, 2.1) for PM_2.5_ and CO, respectively.

**Figure 3.**
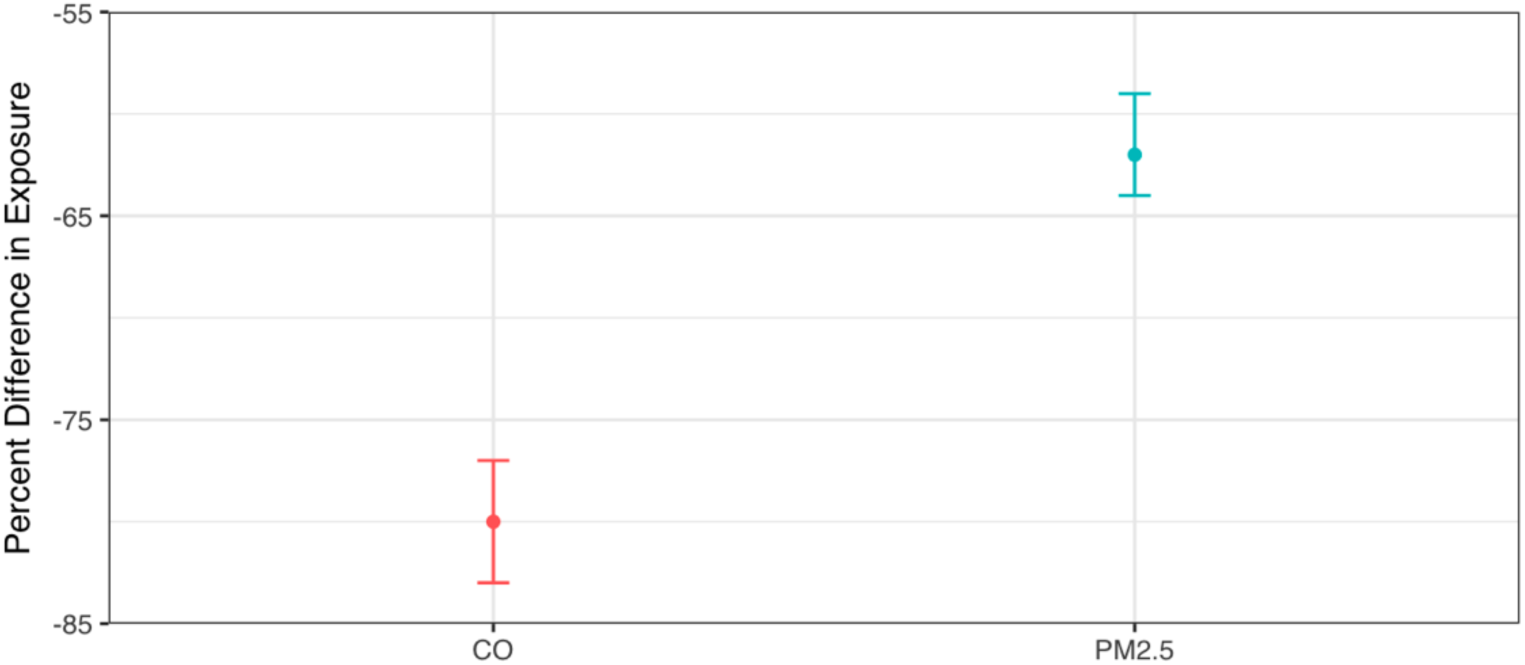
Modeled impacts of the HAPIN LPG intervention on child’s PM_2.5_ and CO exposure. Linear mixed-effects models had log-transformed pollutant exposures as the dependent variable. Whiskers are 95% confidence intervals. Data presented contrast the intervention arm with the control arm. Note: HAPIN, Household Air Pollution Intervention Network; LPG, liquefied petroleum gas; PM_2.5_, particulate matter with an aerodynamic diameter less than or equal to 2.5 µm; CO, carbon monoxide.

## Discussion

Our findings add to evidence of the effectiveness of a well-used and highly adhered to LPG intervention for lowering exposures to household air pollution, measured here as PM_2.5_ and CO.^12, 13^ Our findings are consistent with previous work done as part of the HAPIN trial to characterize exposures during pregnancy; among the same households, we find remarkable consistency between exposure reductions during pregnancy and for young children, as reported here. As we reported previously for pregnant women, these exposures are, to the best of our knowledge, among the most substantial for a household energy intervention to date. Our findings of reduced exposures are consistent and stable across our four low-and middle-income settings.

Comparisons with measurements made during HAPIN are especially relevant, as they provide evidence of stable reductions throughout different phases and measurement regimens of the trial. Prior to the birth of the child, maternal exposure to HAP was measured once at baseline, prior to randomization, and at most twice during pregnancy. Findings from those initial measurements of the intervention’s effectiveness indicated similar reductions in PM_2.5_ and CO exposures (61% and 81%, respectively, as compared to 62% and 80% in the current study, **Figure 4**). Taken in total, these findings suggest the HAPIN intervention reduced exposures in a significant and consistent pattern throughout pregnancy and the child’s first year of life.

**Figure 4.**
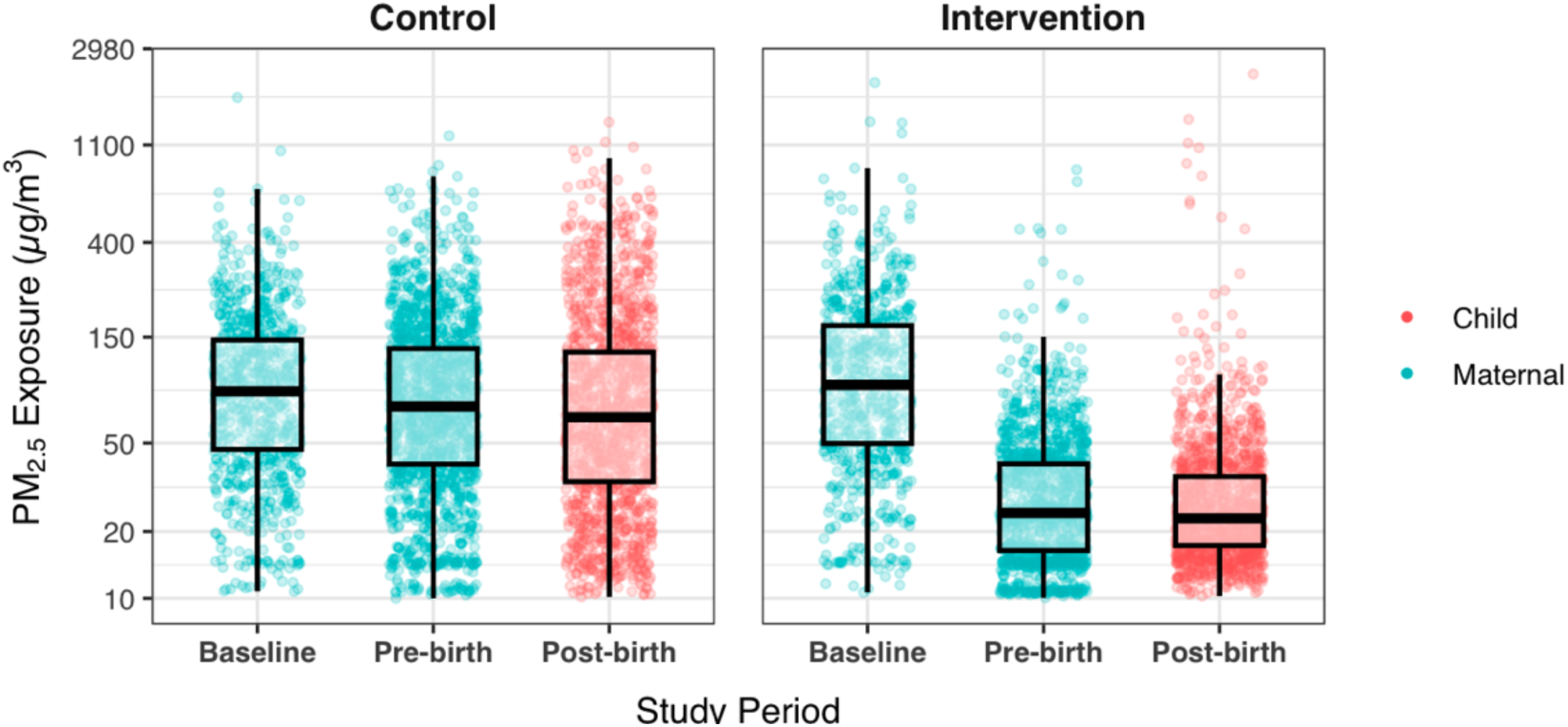
Comparison of PM_2.5_ exposures between maternal measurements made during pregnancy and measurement reconstructed for children post-birth. The *Control* panel contains data for households randomized to continue use of biomass; data in the *Intervention* panel are from households randomized to LPG. Baseline measurements were made prior to randomization. Pre-birth measurements were made during pregnancy. Blue dots indicate maternal measurements previously reported; red dots are findings from this study. Data are presented as box plots. The lower and upper hinges correspond to the first and third quartiles (the 25th and 75th percentiles). The upper and lower whiskers extend 1.5 times the interquartile range above and below the upper and lower hinges. Data beyond the whiskers are outliers.

Several studies have measured personal exposure to HAP among children under five. Early studies in Guatemala,^30, 31^ Gambia,^32–34^ and South Africa^35^ measured CO using passive diffusion tubes attached to children’s clothing. Recent studies used personal monitors to measure children’s exposures to PM_2.5_ in Malawi,^36^ the United States,^37^ Rwanda,^38^ Ghana,^39^ Vietnam,^40^ and CO in Ghana,^8, 17^ Peru,^41^ Rwanda,^42^ or both Uganda and Ethiopia.^43^ Among identified studies, four measured PM_2.5_/CO exposures using personal monitors in children under 1-year old. In these studies, sampling instruments were all attached to the customized clothing of the caregiver.^8, 17, 36, 41, 42^ Several direct measurements were conducted among children between ages 1 and 5 with lightweight personal monitors in a backpack/shoulder bag or a vest pocket.^37, 38, 40^ Studies also used microenvironment measurements plus time-activity logs and statistical modeling to estimate the exposures of children.^32, 39, 43^

Despite these different approaches, reported values are uniformly high among LMIC households using solid fuels. Kirby et al. (2019) reported high child PM_2.5_ exposure in Rwanda, where the median (IQR) exposure was 154 (85–267) µg/m^3^ in the intervention households using an improved biomass stove and 161 (91–285) µg/m^3^ in control households.^38^ In Malawi, the geometric mean (GM) (95% CI) of PM_2.5_ exposure among children at six months was 53.9 (49.4 - 58.7) in a cleaner-burning biomass-fueled cookstove intervention group and 49.0 (45.0 - 53.3) in the control group using open fires.^36^ Similarly, the GM (GSD) of young infants’ exposure to PM_2.5_ in households using biomass was 80.2 (1.34) and 97.0 (1.89) in Uganda and Ethiopia, respectively.^43^ Clean fuels resulted in lower exposures; in Vietnam, the reported mean (SD) child exposure was 36 (24) µg/m^3^ in households using electricity for cooking and 44 (38) µg/m^3^ in homes using LPG.^40^

Personal CO exposure of children under five old followed a similar pattern. The 48-hour median (IQR) personal exposure observed in Ghana was 1) 0.48 (0.17–1.23) ppm in traditional cookstove households, 2) 0.51 (0.17–1.10) ppm in improved biomass cookstove households, and 3) 0.39 (0.12–0.94) ppm in LPG stove households.^8^ In Rwanda, the estimated child CO exposure from burning solid fuels was much higher, with a 24-hour mean of 6.8 ppm.^42^ The reported 24-hour GM was 1.4 in Peru,^41^ 0.64 in Uganda, and 2.4 in Ethiopia.^43^

Our study has a number of strengths. It contributes to this growing body of evidence of HAP exposures among young children. It is among a small number that has used sensor-based measurements to assess microenvironmental locations and coupled this time-activity data with microenvironmental PM_2.5_ and CO measures, enabling exposure reconstruction. We also acknowledge a number of limitations. While we highlight the reduced exposures for both expecting mothers and their young children resulting from the HAPIN intervention, we acknowledge fairly large, unexplained residual exposures. Future work should focus on identifying the drivers of this residual exposure, which may include outdoor exposures from trash burning, from unmonitored biomass stoves, or from other household energy sources (animal food preparation, heating water for bathing, etc). Second, we acknowledge the unique circumstances of the trial, in which LPG was supplied for free, and during which households were provided behavioral nudges to continue its use, neither of which will continue post-trial. Third, despite the scale of the exposure assessment exercise reported here, we only reconstructed periodic snapshots of exposure that may not represent the ‘true’ participant experience. However, we note the remarkable stability of both the control arm and intervention exposures post-randomization (**Figure 4**) for mothers and children. Next, microenvironmental reconstructions required the use of numerous data streams linked by unique beacons, beacon loggers, and household IDs. Loss of any single data stream – from data processing errors, file naming errors, or mis-entry during digital record taking while deploying samples – resulted in lost samples. Future work will describe efforts to model both mother and children’s exposure using limited subsets of the data collected during HAPIN and leveraging repeat measurements in the same household, where available. Finally, we acknowledge that the study children’s behavioral patterns at the three measurement timepoints likely are not representative of each other nor of later periods of childhood and adolescence. Nonetheless, the findings provide evidence (and rationale) for these types of measurements and for further investigation of exposures during critical periods of child development.

While the HAPIN study is an efficacy trial under which LPG was provided continuously and for free, our findings, coupled with previous findings for pregnant women^14^ and forthcoming findings for non-pregnant adult women from the same households (*in preparation*), provide evidence that adoption and sustained, near-exclusive use of clean fuels can reduce exposure to household air pollution for multiple household members, including vulnerable young children. Given the potentially deleterious impacts of exposure across the lifespan, additional measurements are warranted for children and other household members whose experience of pollution is not currently well characterized.

## Data Availability

All data produced in the present study are available upon reasonable request to the authors

## Acknowledgements

The HAPIN trial is funded by the U.S. National Institutes of Health (cooperative agreement <u>1UM1HL134590</u>) in collaboration with the Bill & Melinda Gates Foundation [OPP1131279].

The investigators would like to thank the members of the advisory committee - Patrick Breysse, Donna Spiegelman, and Joel Kaufman - for their valuable insight and guidance throughout the implementation of the trial. We also wish to acknowledge all research staff and study participants for their dedication to and participation in this important trial.

A multidisciplinary, independent Data and Safety Monitoring Board (DSMB) appointed by the National Heart, Lung, and Blood Institute (NHLBI) monitors the quality of the data and protects the safety of patients enrolled in the HAPIN trial. NHLBI DSMB: Nancy R. Cook, Stephen Hecht, Catherine Karr (Chair), Joseph Millum, Nalini Sathiakumar, Paul K. Whelton, and Gail Weinmann and Thomas Croxton (Executive Secretaries). Program Coordination: Gail Rodgers, Bill & Melinda Gates Foundation; Claudia L. Thompson, National Institute of Environmental Health Sciences; Mark J. Parascandola, National Cancer Institute; Marion Koso-Thomas, Eunice Kennedy Shriver National Institute of Child Health and Human Development; Joshua P. Rosenthal, Fogarty International Center; Concepcion R. Nierras, NIH Office of Strategic Coordination Common Fund; Katherine Kavounis, Dong-Yun Kim, Antonello Punturieri, and Barry S. Schmetter, NHLBI.

The findings and conclusions in this report are those of the authors and do not necessarily represent the official position of the US National Institutes of Health or Department of Health and Human Services.

## Notes

### Competing Interest Statement

The authors have declared no competing interest.

### Clinical Trial

NCT02944682

### Funding Statement

This study was funded by the U.S. National Institutes of Health (NIH; cooperative agreement 1UM1HL134590) in collaboration with the Bill & Melinda Gates Foundation (OPP1131279).

### Author Declarations

The study protocol has been reviewed and approved by institutional review boards (IRBs) and Ethics Committees at Emory University (00089799), Johns Hopkins University (00007403), Sri Ramachandra Institute of Higher Education and Research (IEC-N1/16/JUL/54/49), the Indian Council of Medical Research - Health Ministry Screening Committee (5/8/4-30/(Env)/Indo-US/2016-NCD-I), Universidad del Valle de Guatemala (146-08-2016), Guatemalan Ministry of Health National Ethics Committee (11-2016), Asociacion Benefica PRISMA (CE2981.17), the London School of Hygiene and Tropical Medicine (11664-5), the Rwandan National Ethics Committee (No.357/RNEC/2018), and Washington University in St. Louis (201611159). The study has been registered with ClinicalTrials.gov (Identifier NCT02944682).

