## Supplemental Information for "Post-birth exposure contrasts for children during the Household Air Pollution Intervention Network randomized controlled trial"

**Table S1**. Overall indirect $PM_{2.5}$ exposure data completeness of children in HAPIN Trial by IRC and visit

| IRC | Visit | Enrolled | Exposure visits | Exposure data | Fraction with valid exposure data (%) | Fraction of exposure visits with valid data (%) |
|---|---|---|---|---|---|---|
| Guatemala | B1 | 754 | 678 | 514 | 68.2 | 75.8 |
| | B2 | 748 | 638 | 404 | 54 | 63.3 |
| | B4 | 744 | 689 | 284 | 38.2 | 41.2 |
| India | B1 | 761 | 672 | 285 | 37.5 | 42.4 |
| | B2 | 761 | 684 | 229 | 30.1 | 33.5 |
| | B4 | 758 | 724 | 230 | 30.3 | 31.8 |
| Peru | B1 | 722 | 417 | 172 | 23.8 | 41.2 |
| | B2 | 715 | 406 | 138 | 19.3 | 34 |
| | B4 | 711 | 520 | 129 | 18.1 | 24.8 |
| Rwanda | B1 | 749 | 602 | 214 | 28.6 | 35.5 |
| | B2 | 745 | 614 | 148 | 19.9 | 24.1 |
| | B4 | 736 | 583 | 33 | 4.5 | 5.7 |

**Table S2**. Overall indirect $PM_{2.5}$ exposure data completeness of children in HAPIN Trial by IRC, visit, and study arm

| IRC | Visit | Arm | Enrolled | Exposure visits | Exposure data | Fraction with valid exposure data (%) | Fraction of exposure visits with valid data (%) |
|---|---|---|---|---|---|---|---|
| Guatemala | B1 | Control | 378 | 337 | 259 | 68.5 | 76.9 |
| | | Intervention | 376 | 341 | 255 | 67.8 | 74.8 |
| | B2 | Control | 374 | 318 | 200 | 53.5 | 62.9 |
| | | Intervention | 374 | 320 | 204 | 54.5 | 63.8 |
| | B4 | Control | 373 | 351 | 133 | 35.7 | 37.9 |
| | | Intervention | 371 | 338 | 151 | 40.7 | 44.7 |
| India | B1 | Control | 380 | 330 | 183 | 48.2 | 55.5 |
| | | Intervention | 381 | 342 | 102 | 26.8 | 29.8 |
| | B2 | Control | 380 | 343 | 146 | 38.4 | 42.6 |
| | | Intervention | 381 | 341 | 83 | 21.8 | 24.3 |
| | B4 | Control | 378 | 361 | 149 | 39.4 | 41.3 |
| | | Intervention | 380 | 363 | 81 | 21.3 | 22.3 |
| Peru | B1 | Control | 347 | 196 | 75 | 21.6 | 38.3 |
| | | Intervention | 375 | 221 | 97 | 25.9 | 43.9 |
| | B2 | Control | 341 | 191 | 59 | 17.3 | 30.9 |
| | | Intervention | 374 | 215 | 79 | 21.1 | 36.7 |
| | B4 | Control | 339 | 250 | 59 | 17.4 | 23.6 |
| | | Intervention | 372 | 270 | 70 | 18.8 | 25.9 |
| Rwanda | B1 | Control | 384 | 310 | 112 | 29.2 | 36.1 |
| | | Intervention | 365 | 292 | 102 | 27.9 | 34.9 |
| | B2 | Control | 382 | 310 | 60 | 15.7 | 19.4 |
| | | Intervention | 363 | 304 | 88 | 24.2 | 28.9 |
| | B4 | Control | 379 | 306 | 17 | 4.5 | 5.6 |
| | | Intervention | 357 | 277 | 16 | 4.5 | 5.8 |

**Table S3**. Overall indirect CO exposure data completeness of children in HAPIN Trial by IRC and visit

| IRC | Visit | Enrolled | Exposure visits | Exposure data | Fraction with valid exposure data (%) | Fraction of exposure visits with valid data (%) |
|---|---|---|---|---|---|---|
| Guatemala | B1 | 754 | 678 | 486 | 64.5 | 71.7 |
| | B2 | 748 | 638 | 387 | 51.7 | 60.7 |
| | B4 | 744 | 689 | 281 | 37.8 | 40.8 |
| India | B1 | 761 | 672 | 362 | 47.6 | 53.9 |
| | B2 | 761 | 684 | 310 | 40.7 | 45.3 |
| | B4 | 758 | 724 | 295 | 38.9 | 40.7 |
| Peru | B1 | 722 | 417 | 163 | 22.6 | 39.1 |
| | B2 | 715 | 406 | 135 | 18.9 | 33.3 |
| | B4 | 711 | 520 | 94 | 13.2 | 18.1 |
| Rwanda | B1 | 749 | 602 | 213 | 28.4 | 35.4 |
| | B2 | 745 | 614 | 143 | 19.2 | 23.3 |
| | B4 | 736 | 583 | 37 | 5 | 6.3 |

**Table S4**. Overall indirect CO exposure data completeness of children in HAPIN Trial by IRC, visit, and study arm

| IRC | Visit | Arm | Enrolled | Exposure visits | Exposure data | Fraction with valid exposure data (%) | Fraction of exposure visits with valid data (%) |
|---|---|---|---|---|---|---|---|
| Guatemala | B1 | Control | 378 | 337 | 248 | 65.6 | 73.6 |
| | | Intervention | 376 | 341 | 238 | 63.3 | 69.8 |
| | B2 | Control | 374 | 318 | 190 | 50.8 | 59.7 |
| | | Intervention | 374 | 320 | 197 | 52.7 | 61.6 |
| | B4 | Control | 373 | 351 | 134 | 35.9 | 38.2 |
| | | Intervention | 371 | 338 | 147 | 39.6 | 43.5 |
| India | B1 | Control | 380 | 330 | 177 | 46.6 | 53.6 |
| | | Intervention | 381 | 342 | 185 | 48.6 | 54.1 |
| | B2 | Control | 380 | 343 | 151 | 39.7 | 44 |
| | | Intervention | 381 | 341 | 159 | 41.7 | 46.6 |
| | B4 | Control | 378 | 361 | 142 | 37.6 | 39.3 |
| | | Intervention | 380 | 363 | 153 | 40.3 | 42.1 |
| Peru | B1 | Control | 347 | 196 | 68 | 19.6 | 34.7 |
| | | Intervention | 375 | 221 | 95 | 25.3 | 43 |
| | B2 | Control | 341 | 191 | 59 | 17.3 | 30.9 |
| | | Intervention | 374 | 215 | 76 | 20.3 | 35.3 |
| | B4 | Control | 339 | 250 | 48 | 14.2 | 19.2 |
| | | Intervention | 372 | 270 | 46 | 12.4 | 17 |
| Rwanda | B1 | Control | 384 | 310 | 111 | 28.9 | 35.8 |
| | | Intervention | 365 | 292 | 102 | 27.9 | 34.9 |
| | B2 | Control | 382 | 310 | 59 | 15.4 | 19 |
| | | Intervention | 363 | 304 | 84 | 23.1 | 27.6 |
| | B4 | Control | 379 | 306 | 17 | 4.5 | 5.6 |
| | | Intervention | 357 | 277 | 20 | 5.6 | 7.2 |

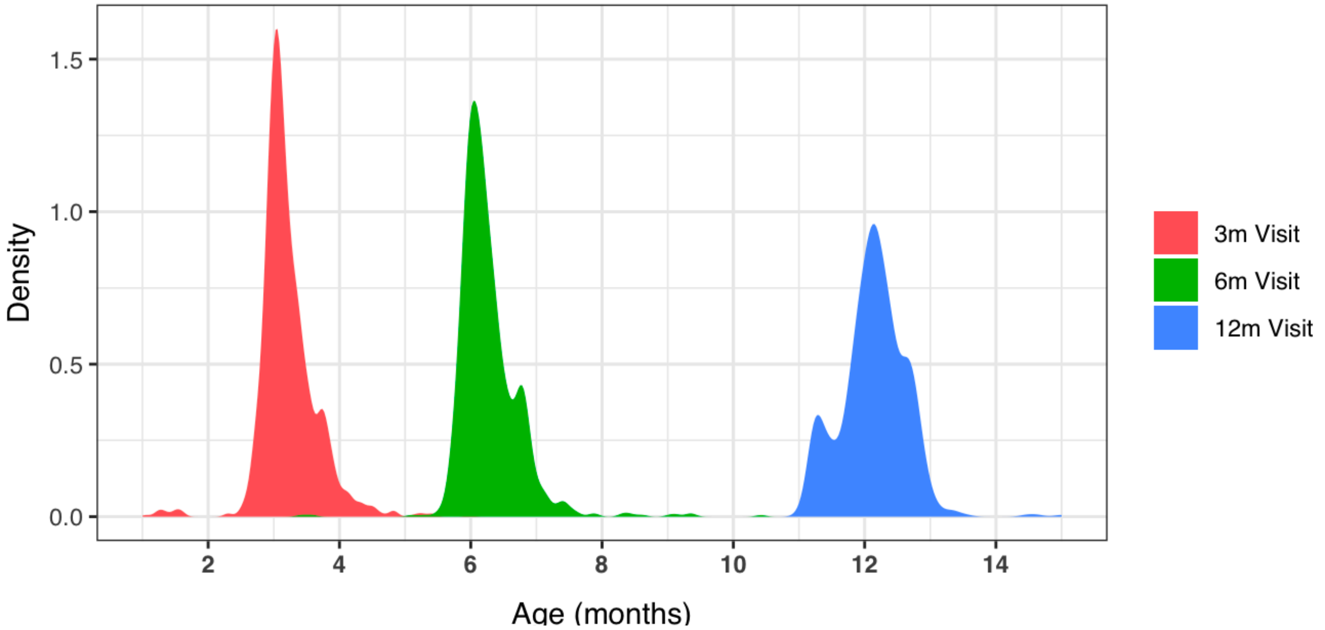


**Figure S1**. Distributions of child's age in months at each study visit.

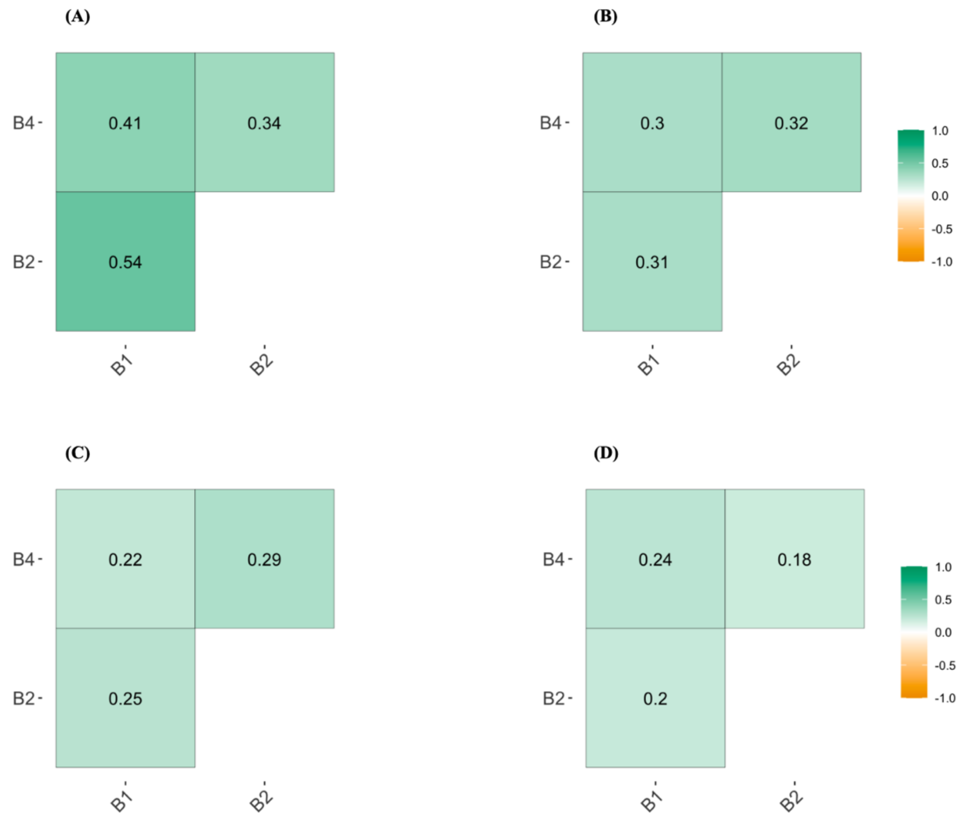


**Figure S2**. Correlations of measured $PM_{2.5}$ (A and B) and CO (C and D) exposures by study visit for control (A and C) and intervention households (B and D). B1 indicates visits at <3m, B2 indicates visits at ~6m, and B4 indicates visits at approximately 12m.
